# Iterative Development of a Mobile Phone Application to Support Community Health Volunteers during Cervical Cancer Screening in Western Kenya

**DOI:** 10.1101/2020.09.23.20200162

**Authors:** Jacob Stocks, Yujung Choi, Ibrahim Saduma, Megan Huchko

## Abstract

**Background:** To achieve the World Health Organization targets for cervical cancer elimination, low- and middle income countries must develop innovative strategies to provide human papillomavirus (HPV) -based screening at a population level. Mobile health may help fill gaps in electronic specimen tracking and patient education. We sought to develop a mobile health application (“mSaada”) to support HPV-based screening in partnership with community health volunteers and program planners in western Kenya.

**Methods:** A team of student programmers developed a prototype to meet previously identified gaps in screening patient education, protocol support, data capture and specimen tracking. The protoype was iteratively developed through three waves of in-person working sessions with quantitative and qualitative feedback, with planned improvements in mSaada functionality after each wave of in-person data collection. Results: Twelve Community Health Volunteers (CHVs) and clinicians took part in the in-person sessions. Participants found mSaada useful and easy to use. Key feedback was used to alter the appearance of the mainframes, add translation in additional local languages and change potentially embarrassing figures. Participants also suggested workflow design and technology needs necessary for sustainability.

**Conclusion:** Using a process of iterative feedback with key stakeholders and rapid response from developers, we have developed a mobile application ready for pilot testing in HPV-based screening programs led by CHVs.

## Background

According to projections from the World Health Organization (WHO), incident cases of and annual deaths due to cervical cancer could increase by 37% and 47% to 777,000 and 459,000, respectively, by 2040.^1^ The majority of this disease burden is observed in low- and middle-income countries (LMICs) in Sub-Sharan Africa (SSA) and South East Asia.^1,2^ In order to reduce rates of morbidity and mortality due to cervical cancer, high quality screening programs which reach the target population in a cost-effective and culturally appropriate manner are needed. While cervical cancer-related morbidity and mortality are highly preventable with timely and effective screening and treatment,^3^ such programs are nascent in many LMICs and face infrastructural and financial challenges.^4,5^ Kenya’s Ministry of Health (MoH) has developed national guidelines for cervical cancer screening, which align with the WHO-endorsed framework for human papillomavirus (HPV) based screening. While this may drive greater access to services throughout the country, there are significant barriers to implementation.^3^ One such barrier is the use of non-formally trained lay-providers, such as community health volunteers (CHVs), to deliver care. While these individuals can reduce the clinical load on an already strained health system, they require adequate supervision and support to carry out service delivery.

Poor adherence to clinical guidelines has been cited as a challenge to utilizing CHV-led programs and a risk of delivering substandard care. A CARE-led initiative to employ lay health providers in Siaya district, Kenya showed poor adherence to clinical guidelines by trained community health workers.^6^ Further training, on-the-job supervision, and technical support have been shown to increase quality of service delivery. One study conducted in the Morogoro region of Tanzania found that lay health workers “value supervision and appreciate the sense of legitimacy that arises when supervisors visit them in the village.”^7^ However, it is important that when supervision is offered, it occurs in a respectful, non-judgmental way such that CHVs do not feel criticized or embarrassed of their work. Such poor-quality supervision could lead to decreased self-efficacy and less desire to complete assigned tasks. The provision of aids, such as pamphlets, flipcharts, mobile applications, and handbooks have been cited as effective at increasing adherence and may provide a means to avoid overly critical supervision that still yields the necessary support.^6,8^

Mobile health (mhealth) has been defined by the WHO as “medical and public health practice supported by mobile devices, such as mobile phones, patient monitoring devices, personal digital assistants, and other wireless devices.”^9^ mHealth approaches, which typically employ information sharing, education, communication, or data collection strategies in order to meet specified goals,^10^ have been used to deliver service reminders, promote behavior change, enhance medication adherence, provide health education, and collect and store patient data.^11–15^ There is a large body of evidence supporting the feasibility, acceptability, and usability of a variety of mhealth interventions that target a wide range of health conditions such as depression, diabetes, HIV/AIDS, and cancer.^16–22^ While many mhealth interventions are patient-sided, there are a growing number of provider-sided applications which seek to support and improve the delivery of health services.^10,16^ These novel approaches provide a platform for the effective delivery of evidence-based practices targeting a range of health outcomes in often unreached or hard-to-reach populations, and can be used to support task shifting of health delivery to CHVs or other lay health workers.^23^

Effective implementation of mhealth interventions is reliant upon appropriate, context-specific development that is acceptable and useable by the target population.^24,25^ Thus, mhealth interventions should be designed such that they are context specific and reflective of mobile phone ownership and rates of usage. Such considerations must be incorporated in the design and development stages of mobile applications and other interventions in order to yield final products which reflect the needs, desires, and capabilities of targets users.^25^ Otherwise, low levels of usefulness and user uptake could result from poor development.^26^ Recent work by Huchko et al. showed that CHVs delivering cervical cancer screening in community- and facility-based settings in western Kenya desired more protocol and decision support tools in order to effectively complete screening.^27^ In addition, the study identified a clear need for continued education regarding the cause, risk, transmission, and prevention of Human Papillomavirus (HPV).^28–30^ Based on high reported mobile phone ownership within Kenya and past research documenting success with text message-based delivery of screening results in western Kenya,^31,32^ the introduction of a mobile application-based intervention to address gaps in education and provider support appears feasible in this context. The aim of this study was to iteratively develop and refine the *mSaada* mobile application in consultation with key stakeholders prior to small-scale pilot testing in health facilities in western Kenya.

## Methods

This two-wave qualitative study gathered detailed insight into perspectives on the functionality and use of a newly developed mobile phone application. Data collection for the present study took place in Migori and Kisumu, Kenya. Both locations, one rural and one urban, maintain existing cervical cancer screening programs within local health facilities. However, service uptake is low among women of reproductive age. We chose these locations based on the target end-user of the application, community health volunteers, which are commonly employed in both places.

mSaada, meaning “support” in Kiswahili, is a counseling and decision support tool designed for use by CHVs. As an android-based mobile phone application developed by students as part of a computer science course at Duke University, mSaada was designed to address logistical and educational gaps in HPV-based cervical cancer screening in western Kenya. The app features were developed based on feedback from prior research in the team, and through virtual communication with Kenyan key stakeholders, who provided feedback on the overall functionality, wireframes and pilot app. As described in Table 1, the app includes four main features to take the CHVs through the entire process of screening, including counseling and decision-making, answers to frequently asked questions, data collection and specimen tracking.

**Table 1.**
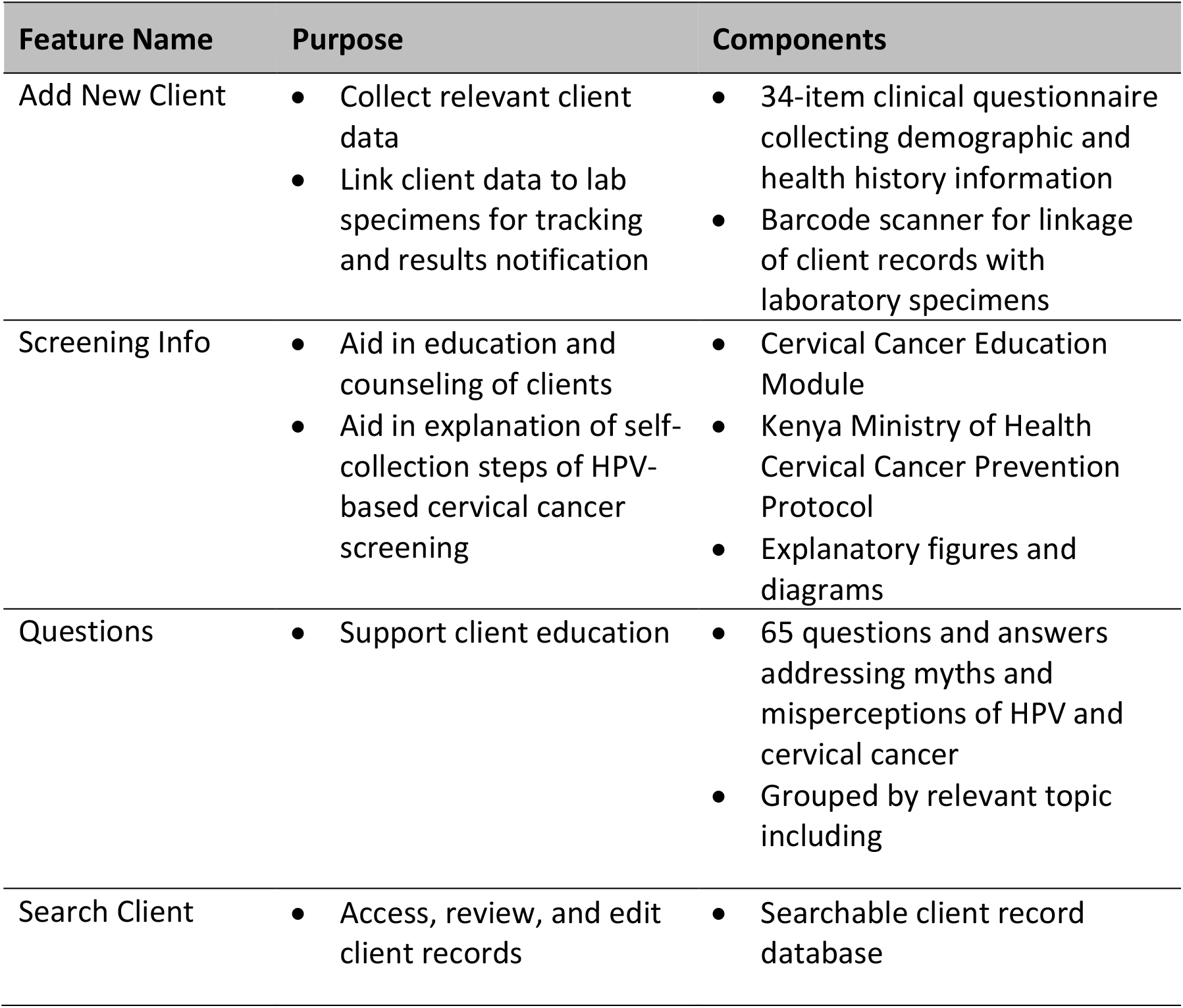
Description of mSaada mobile application features.

To gather a wide variety of input and perspectives regarding mSaada and its features, we recruited individuals in three categories: experts (n=6), end-users (n=6), and lay-persons (n=6). Expert study participants were individuals with direct experience providing cervical cancer related services, such as clinicians or study staff working in related research. End-user study participants were individuals who had worked or were currently working as CHVs and had performed cervical cancer screening using HPV testing via self-collection. Lay-person study participants were individuals who had no formal training or experience in cervical cancer screening. Research staff identified potential study participants based on prior engagement with the individuals or by recommendation from partner facility staff. Study staff recruited participants by phone or in-person prior to the beginning of the study period.

The iterative development period lasted eight weeks in total and consisted of alternating waves of data collection and integration of feedback into the mSaada platform (Figure 1). During data collection waves, participants attended day-long feedback sessions which were categorized by participant group (i.e. experts, end-users, and lay-persons). To obtain a combination of novel and continuous feedback on mSaada, we asked two representative experts, end-users, and lay-persons (n=6) to participate in both waves of data collection activities, completing an in-depth interview during both waves. All other study participants (n=12) were asked to attend only one day of feedback sessions, completing only one in-depth interview. This resulted in a total of 24 in-depth interviews between 18 participants, with an average length of 45 minutes.

**Figure 1.**
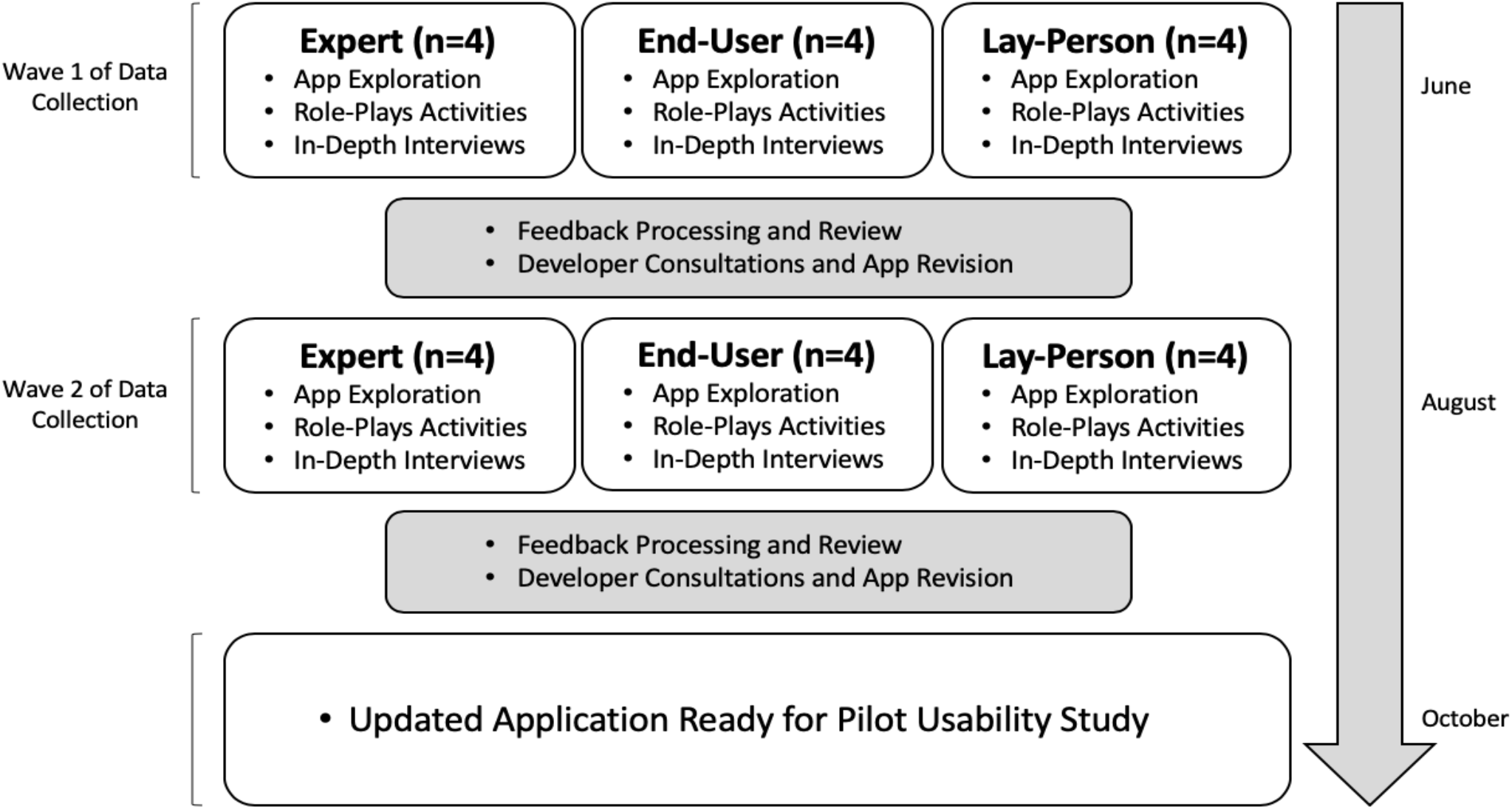
Iterative development timeline and activities.

Feedback sessions (n=6) began with a description of study aims, intended methods of data collection, and completion of written informed consent. Following this discussion, researchers provided detailed, screen-by-screen explanations of the mSaada app and its features, allowing participants to follow along using study phones loaded with the platform during the demonstration. After initial hands-on familiarization in a group setting, participants completed simulation activities in pairs, including CHV/Client roleplays and example scenarios to gain more experience with the platform.^33^ Explanation, familiarization, and role-plays with mSaada lasted three hours, on average. Thereafter, we conducted individual in-depth interviews to gather detailed feedback about the platform. Both the group exploration/familiarization sessions and individual in-depth interviews were conducted in English and audio-recorded for transcription. We completed transcription using Otter.ai, a free, open source software. Study staff transcribed recordings using the software and reviewed and edited resulting transcripts for accuracy. Researchers followed the same protocol for both waves of data collection. All study activities occurred within the Kisumu Office of the Duke Center for Global Reproductive Health and the Migori County Hospital. Group feedback sessions were conducted within the conference area of the office while individual in-depth interviews took place within private rooms. Research staff and app developers convened to discuss and integrate key participant feedback after it was gathered. Researchers consolidated data from the first wave of interviews into a master list of recommended changes and updates. The resulting 12-page document was discussed amongst Kisumu- and Durham-based researchers in order to focus and prioritize revision efforts. While most of the recommended changes were considered for integration prior to the second wave of interviews, some participants suggested changes that were not feasible to undertake between the two feedback waves. We addressed these changes, including a large expansion in capabilities of the existing Search Client feature, after the iterative development period.

After agreed upon prioritization of the master list, researchers provided participant feedback to the Nairobi-based app developer for integration. In order to track and discuss progress during the 3-week period of app refinement, the app-developer supplied Kisumu-based researchers with intermittent versions of mSaada, which were downloaded to study phones and reviewed for accuracy. This back-and-forth process helped to facilitate effective communication between team members and allowed for successful completion of app refinement.

We developed a two-part interview guide that was used for all participants during both waves of data collection. First, we asked participants to reflect on each of the four features of the app. For each feature, participants were asked their opinion on usability, user control, aesthetics, comfort, ease of use, and about any challenges they faced. The interview concluded with questions regarding overall impressions of the platform and thoughts about implementation and use of mSaada within a Kenyan context. We asked participants for any recommended changes or updates to the app’s layout and features, and any concerns they had about the app’s use within facilities.

We developed a codebook based on the interview guide and informed by the literature. We analyzed the qualitative data using thematic analysis and a four-stage process.^34^ Analysis was aided by NVivo 12. First, researchers reviewed all transcripts and created document memos to summarize key points from each participant and to get a strong sense of the collected data. Second, we identified deductive structural codes based on the two sections of the interview guide (i.e. feature-specific feedback and overall impressions).

Third, inductive thematic codes were identified via thorough review and re-review of participant transcripts. As thematic codes were identified, researchers added them to the a priori codebook and recoded transcripts based on observed themes. Fourth, after completion of thematic coding, we wrote analytic memos for each identified theme detailing the similarities and differences in feedback between features of mSaada to broadly summarize the gathered information. Interim analysis, completed between waves 1 and 2 of data collection, utilized a similar process but was not aided by NVivo 12. In order to accelerate the procedure, researchers completed a consolidated version of the aforementioned four-stage process.

We obtained ethical approval from Duke University Campus Institutional Review Board (2019-0650). All participants provided written informed consent prior to initiation of study activities.

## Results

Table 2 outlines key updates made to each feature of mSaada. Figure 2 depicts wireframes of the final version of mSaada after iterative development. Qualitative data gathered during the iterative development process was classified into four categories: ease of use, accessibility of information, workflow considerations, and acceptability.

**Table 2.** Feature-specific feedback and the resulting revisions to mSaada.

| **App Feature** | **Weakness Identified** | **Changes/Update Made** |
| --- | --- | --- |
| Add New Client | • Lack of clarity in questionnaire items<br>• Lack of questionnaire items gathering administrative details<br>• Questionnaire items only presented in English | • Refined phrasing in consultation with Kisumu-based staff<br>• Added items regarding screening location and provider name<br>• Translated feature content into Kiswahili and Dholuo |
| Screening Info | • Lack of sufficiently detailed graphics and instructions for self-collection<br>• Need for additional images/graphics to aid in counseling<br>• Content only presented in English | • Updated step-by-step instructions to include more detail<br>• Included Kenya MOH Cervical Cancer Prevention Protocol for additional support<br>• Translated feature content into Kiswahili and Dholuo |
| Questions | • Difficulty using built-in keyword search query<br>• Educational content only presented in English | • Broaden keyword search query to include synonymous terms<br>• Translated into Kiswahili and Dholuo |
| Search Client | • Unable to effectively edit client records<br>• Search fields of database query returning broad group of client records | • Incorporated editing capabilities within feature<br>• Added additional search fields to aid in narrowing of client records |

**Figure 2.**
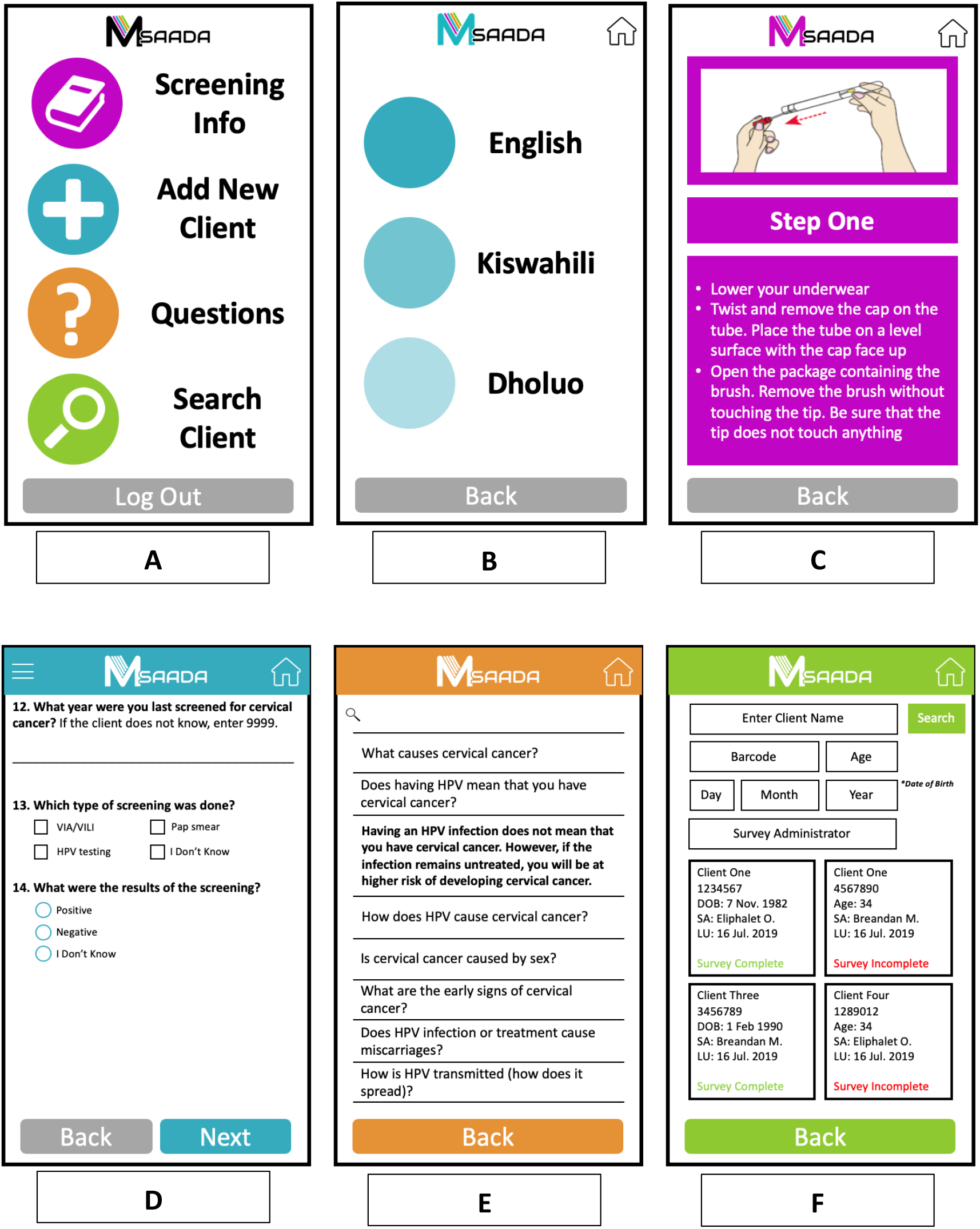
Example wireframes of mSaada: a) homepage depicting four main features; b) page showing option to select language preference; c) self-collection screening instructions within the Screening info feature; d) patient data entry page within the Add New Patient feature; e) searchable frequently asked questions with corresponding answer; f) Search Client feature illustrating client record lookup.

### Ease of Use

Overall, participants were comfortable using the main features of mSaada. Many participants cited previous mobile phone use, specifically smart phone use, as a main reason for their comfort with the app.

> *“The practitioners here in Kisumu, I think most of them are used to these phones, so I don’t think they will have many challenges, maybe it is just a matter of familiarizing with the app. It’s quite different but almost everyone has smartphones, so they’re used to them*.*” (Expert 4)*.

When asked about their use of specific features of mSaada, participants reported ease with all features except the Questions feature. Many applauded the logical flow from screen-to-screen within each feature allowing for overall easy use of the app. In addition, participants liked the use of “Back” and “Next” buttons for navigation within the app and felt the scrolling and swiping aspects of the platform were effective, responsive, and straightforward. Regarding the Questions feature, participants reported difficulty when using the keyword search to look for specific content, finding it difficult to identify the correct keyword necessary to locate the needed information. When asked about their challenges with the Questions feature, participants believed this difficulty was due to a lack of familiarity and experience with mSaada, not a weakness in the design or functionality of the platform. Most participants felt the use of the Questions feature would improve with repeated exposure and did not recommend any changes.

> *“I think the feature helps a lot in replying to questions. All we have to do is master the concept, so that during the [keyword search] you already have an insight. That way when a client asks you a question, you know what to type and then get the correct information*.*” (End-User 4)*

Participants concluded the most effective way to become “well-versed” with the app was through role-play and other simulation activates. Many recommended detailed and interactive trainings for CHVs to facilitate successful use prior to “going to the field.”

### Accessibility of Information

Accessibility of information emerged as both a strength and weakness of mSaada. Overall, participants commended the use of “very simple”, direct, non-medical jargon within the app. Participants suggested this would likely increase the usefulness of mSaada, as CHVs would be able to effectively communicate necessary information with clients at a contextually appropriate level of understanding, regardless of their lack of formal medical training. In addition, the format of educational content, specifically within the Cervical Cancer Education Module, was well received, as participants found the scrollable nature of the electronic information easier to use in comparison with the bulky flip charts currently utilized within facilities.

While strengths in presentation of information were identified, all participants highly recommended translation of app content into Kiswahili, a national language of Kenya, as well as Dholuo, a widely used local language in Western Kenya. Participants felt this translation was necessary so that community health volunteers could effectively convey important information to clients of any language preference. In addition, it was mentioned that, by only presenting an English version of the app, CHVs were responsible for translation of information “on the fly” to non-English speaking clients, which could allow for inconsistencies or inaccuracies in the transmission of information from app to CHV to client. Thus, in order to lower the burden of responsibility on CHVs, participants recommended all information intended for the client consumption be “uniformly” translated to both local languages.

In addition to the inclusion of local languages, many participants highlighted sentences and phrases included within the app’s Add New Client and Screening Info features that were not well understood or did not appropriately convey the intended meaning of the statement within a Kenyan context. In order to ensure effective use of mSaada and reduce varied or unintended interpretation of information, participants placed emphasis on the need to assure phrasing of statements was nuanced and reflective of the speech patterns of those in the target area.

Finally, participants were concerned with the lack of ability to access or edit completed client records once information was entered into the app. Many felt it was important to include an editing aspect within the Search Client feature as well as a way of uniquely identifying client records in order to get “all of the info about [a] person.” Participants cited examples from past experience where small typos had been made or where there were questions about a certain client records that needed further evaluation.

### Workflow Considerations

Overall, participants believed mSaada would help accelerate the screening and client data collection processes. Errors and inefficiencies within the app were highlighted and changes were recommended, especially within the Add New Client feature. Participants emphasized the need for incorporation of clinically relevant logic checks throughout the clinical questionnaire. Recommended checks included the following: age, to ensure clients are within the recommended target age range for screening; history of hysterectomy, to ensure no contraindication of screening; and pregnancy status, which is of clinical importance during the treatment of HPV infection or cervical cancer. In addition, participants recommended refinement of response options, specifically the alphabetizing of lengthy dropdown menus, within the clinical questionnaire in order to make the data collection process more efficient.

Participants also emphasized the need to cater the presentation of information within mSaada towards end-users (CHVs) for successful and efficient use within facilities. Many participants felt the wording of items within the clinical questionnaire should be so that CHVs could read the text directly from the app as if they were talking to a client, rather than having to “reframe” the question after reading. During role-play activities, participants often stumbled over questionnaire items not written in this way.

Finally, participants identified a data persistence issue within the Add New Client feature, which they felt would negatively impact CHV workflow within clinical settings. The possibility of data loss and the need for re-entry of client data concerned many participants.

> *“The issue…was that when you go back [to a previous screen using the “Back” button], now you have to again key in the same set of information and this might be a problem if you have a number of patients who are on the line because we may make errors and we may also want to change [answers] without doing away with the whole information*.*” (Lay-Person 2)*

### Acceptability

Most participants believed mSaada included all components and features necessary to aid CHVs in the successful screening of clients. Participants found the Cervical Cancer Education Module within the ‘Screening Info” feature to be comprehensive, but concise. In addition, while not exhaustive, participants found the Questions feature content to be extremely useful for client education and believed the information covered many of the main topics about which a client might inquire. Participants did, however, recommend the four main features on the home screen be reordered. Participants believed presenting the features in chronological order of a client’s screening visit would help to reduce confusion of app users (CHVs) and help to facilitate the visit.

> *“The Add New Client is what we’ll be using mostly for our new clients. It {Screening Info] should be the second because after adding a patient is when you go to the screening info and the education module. And then you go to Questions. And lastly the Search Patient should be the last because maybe you could use that later, after the interview*.*” (End-User 4)*

Finally, some participants recommended the addition of a fifth feature for administrative purposes. The additional feature, a report generating tool, was recommended to enhance mSaada’s usefulness at the facility- and health systems-level. Participants cited a need for up-to-date records on clinical outcomes and services rendered, which is requested by the Ministry of Health.

## Discussion

To our knowledge we are the first to describe the development of an app designed for health workers offering HPV-based cervical cancer screening services in Kenya. The iterative development process, whereby relevant stakeholders with a diversity of perspectives provided input in a cyclical manner, proved to be effective at creating a viable, functioning application for use in small-scale pilot testing. The stakeholder-engaged, iterative process, which has been employed in many studies,^18–20,35,36^ yielded critical insight that could not have been gathered otherwise. For example, many of the word choices and sentence structures included within the platform were considered to be unclear and not likely to be understood by clients during use, even though this language originated from materials provided by Kenyan research staff. While the app was designed to improve the quality of counseling, having end-users test the app revealed additional concerns about the possibility of miscommunication. One example was the English-only format, designed with English-speaking CHVs in mind. Participants brought up concerns about translating health information “on the fly,” prompting the development of translated versions. These lessons learned call to attention the need for highly engaged, locally driven processes of intervention development, even outside of mhealth approaches. Many mhealth interventions are developed with a singular intent and, therefore, use only one approach to achieve their purpose, either through communication, education, data collection, or information sharing.^10^ This method was countered in the present study, as many participants applauded mSaada’s multifaceted nature, offering a comprehensive solution to many of the challenges experienced in the delivery of screening services. This stands as an example of the importance of multidimensional approaches for future mhealth interventions.

A key strength of the present study is its iterative nature. The mSaada platform underwent multiple rounds of testing and refinement in the two-month study period and this process involved a variety of individuals of varying engagement in cervical cancer screening. Also, key stakeholder and end-user feedback was integral to decision-making and revisions of the platform. A study by Fishbein et al., showed the inclusion of a broad range of stakeholders and perspectives proved beneficial for the development of the app, and the sense of co-creation likely contributed to the resulting acceptability of the application.^19^ By striving to produce a product which was context-specific and relevant to its target audience, those with the most first-hand knowledge and deepest insight into the success and failure of current screening efforts were able to drive mSaada’s development, likely resulting in a better and more useful final product. Another strength of this study was the use of qualitative methods to provide detailed feedback, opinions, and perspective on the app’s development and use. Qualitative methods, if executed well, can produce a plethora of actionable information for use in intervention development and further revision. Many usability testing frameworks emphasize the use of qualitative methods when developing and evaluating mhealth interventions.^33^

There were also a few limitations to the present study. First, while we encouraged feedback from a variety of individuals, our small sample size could have missed important feedback. Given the novel nature of the application within this setting, the present study was focused on feedback from application end-users. However, feedback from women being screened by providers using the application is critical to its implementation and was missing from our study. Also, while consulted informally, hospital and district health administrators were not included within our study sample and, therefore, insight into how mSaada might integrate into facilities from a macro-level was not gathered. Finally, participants were only given three hours, on average, to use the application prior to providing feedback. This limited interaction may not have provided sufficient time to fully explore and identify an issue with the platform and functionality in clinical settings should be testing in a pilot study. Future mhealth development studies should strive to gather feedback from a sufficient number of stakeholders at all levels of the health system and provide ample interaction with their proposed platforms.

## Conclusions

This study demonstrates the usefulness of iterative approaches to mhealth development. By engaging a variety of key stakeholders, we were able to quickly develop a mobile app which would be well-received, have ownership among end-users, and ensure readiness for small-scale pilot testing. In the present study, we show a process for an iterative approach to app development that builds on context-specific preliminary work to further improve the functionality prior to introduction in a clinical setting.

## Data Availability

Data is available upon request

